# Incidence of and Household Responses to Pediatric Diarrheal Disease and Acute Respiratory Infections over Time: Protocol for a Cohort Study

**DOI:** 10.64898/2026.08.13.26358877

**Authors:** Emily Treleaven, Indra Chaudhary, Mary Dwan, Rajendra Ghimire, Grace Noppert, John Kubale, Asmita Sharma, Yograj Sharma, Andrew Hashikawa, William G. Axinn, Dirgha J. Ghimire

## Abstract

**Introduction:** Diarrheal diseases and acute respiratory infections (ARI) disproportionately affect young children and families facing disadvantages at the individual, household, and neighborhood level. This unequal burden especially impacts children in low-and middle-income countries, such as Nepal. Data limitations impede the ability to understand children’s illness episodes and treatment trajectories across the course of early childhood and their relationship to household- and neighborhood-level social determinants of health. The Chitwan Valley Family Study (CVFS) is a 30-year panel study providing a wealth of information about household- and neighborhood-level social determinants in Southern Nepal. Drawing on a cohort of young children in CVFS households, this study will measure incidents of acute illness among children under five and leverage existing data from the panel study to understand how intergenerational disadvantages, place, and other social determinants affect the frequency and duration of childhood illness and subsequent healthcare utilization.

**Methods and Analysis:** This study will use daily symptom diaries to track children’s illness symptoms (diarrhea, fever, cough, runny nose, difficulty breathing or wheezing, fatigue, loss of appetite) over the course of a year. Mother respondents will complete a baseline interview, daily symptom diaries, and a weekly phone interview with a trained interviewer to describe the prior week’s symptoms and, in the case of any symptoms, healthcare utilization, treatment, expenditures, and related information. All eligible children aged 3-59 months may participate in two waves of 52 weeks of data collection. We will measure the frequency and duration of diarrhea and ARI, healthcare utilization outcomes, socio-economic status, and distance to healthcare facilities, then merge these measures with prior CVFS data related to parents’ childhood circumstances, health facility characteristics, and neighborhood characteristics.

**Ethics and Dissemination:** We received IRB approval from the Nepal Health Research Council and the University of Michigan. Informed consent will be obtained from respondents for all aspects of data collection. Identifying information will be restricted to the data collection team in Nepal and stored separately from survey data. Interviewers will check that all children with danger signs identified according to WHO/UNICEF Integrated Management of Childhood Illness clinical guidelines have received adequate treatment; a study nurse will follow up and refer those who have not. We will disseminate study findings to respondents, local partners, and nationally in Nepal, as well as in academic journals and at conferences. Datasets will be available for public and restricted-use through the Data Sharing for Demographic Research program at the Inter-university Consortium for Political and Social Research at the University of Michigan.

**Article summary:** *Strengths and limitations of this study:*

- Study respondents will participate in two waves of data collection and complete daily symptom diaries and weekly interviews for 52-weeks for each wave.
- Respondents will record the presence of diarrhea, fever, cough, runny nose, difficulty breathing or wheezing, fatigue, loss of appetite in children aged 3-59 months.
- This study will measure symptoms of acute illness in children under five and will be nested within a 30-year panel study in Nepal.
- We will investigate the relationship between incidence and duration of illness and intergenerational disadvantage, measured using 30-years of existing data.

## INTRODUCTION

Globally, over 2.5 million children under five years of age die each year from infectious diseases that are substantially mitigated with access to basic health-promoting resources such as clean water, vaccination, and timely, quality healthcare [1–3]. The distribution of these diseases, including diarrheal diseases and acute respiratory infections (ARI) such as pneumonia, is not equitable between or within countries [4–6], disproportionately affecting disadvantaged families and communities [7, 8]. Children younger than five years of age are especially vulnerable to infection because of immature immune systems, limited ability to communicate symptoms, and dependence on parents and caregivers [9–11]. Illness during this period of rapid growth and brain development can disrupt nutrition, development, family functioning, and early learning, with potential long-term consequences for health, education, and well-being [12–15]. A broad body of research underscores how social determinants of health including individual- and family-socioeconomic status (SES) and place are important factors in shaping the burden of disease and familial response to acute illness in early childhood [3, 10, 16–23]. These disadvantages vary across domains—including place, wealth, and education—and compound to create significant barriers to health and healthcare for young children and their families [24–29]. Persistent, cumulative disadvantage experienced over multiple generations may have a distinct effect on health and health disparities in early childhood, separate from contemporaneous SES [30–33]. Understanding the underlying social patterning of disparities in pediatric infectious disease is necessary to design and implement effective interventions to reduce morbidity and mortality in early childhood, as well as estimating the cost-effectiveness of such interventions.

Data limitations continue to impede our ability to study the social and biological processes underlying children’s illness episodes and treatment trajectories across the course of early childhood. Studies with rigorous measures of infectious disease in early childhood often lack detailed, time-varying socio-demographic, contextual, or health service characteristics [25, 34–39]; many studies examining healthcare utilization among young children measure SES and/or household, contextual and/or health service characteristics at a single point in time [40–44], hampering analyses of the cumulative effects of these key social determinants of health. Especially rare are multigenerational studies that include prospective measures of parents’ own childhood circumstances that can be linked to their child’s health outcomes. Thus, several important gaps remain, including: (1) how persistent, intergenerational disadvantage—that is, experiences of low SES and related stressors endured by two or more generations in a household—associates with the burden of infectious disease in early childhood; (2) the specific ways in which intergenerational disadvantage and greater burden of disease affect familial responses to acute illness, including healthcare utilization during specific illness episodes and over time, and (3) whether and how place (e.g., distance to health services, concentration of disadvantage) interact with household-level intergenerational disadvantage to shape burden of disease and responses to acute illness.

Previous research suggests there is a complex relationship between household-level socio-economic disadvantage and the burden of infectious disease and mortality in young children [16, 45–55]. For example, a systematic review found that children under five years exposed to indoor air pollution had 45% greater odds of respiratory syncytial virus-associated acute lower respiratory infection compared to unexposed children [48]. Similarly, a study in Indonesia identified a 45% increase in odds of diarrhea for children under five living in households with shared toilet facilities, compared to those with private toilet facilities in low- and middle-income households [47]. We hypothesize that intergenerational disadvantage captures a unique and salient aspect of household disadvantage, the persistent constraint of resources and social capital that can be used to access material resources [56–62], including health care, and that intergenerational disadvantage is associated with higher frequency and greater duration of acute illness episodes.

Beyond the burden of disease, evidence suggests that children who experience more frequent episodes and/or longer duration of illness with diarrhea and/or ARI are more likely to seek care [63, 64]. Yet, because families typically incur both direct and indirect costs for seeking curative health care for acute illnesses for young children [18, 65–67], and seeking health care is often an iterative process [10], a high disease burden may affect disadvantaged and non-disadvantaged families differently. Studies have found that poorer families have relatively greater expenditures on health care for acute illnesses in early childhood [68] and that expenditures for health care for young children can drive families into debt [69, 70].

This study builds on the Chitwan Valley Family Study (CVFS), a 30-year whole family and community panel study in Nepal. The CVFS offers a wealth of data on household and community dynamics over more than three decades, that can be linked to the new data collection in this project. We will leverage new high-frequency data on children’s ARI and diarrheal disease and subsequent healthcare utilization, linked to existing CVFS individual, household, and neighborhood data. In this study, we will examine whether children whose families have faced social and/or economic disadvantage in multiple generations (child, parent, and/or grandparents) are more likely to get sick more frequently and/or for longer than other children, and whether these families respond differently to illness as they make decisions about treatment and healthcare. We focus on two types of common childhood illnesses: diarrheal disease and ARI.

The objective of this study is to analyze household and neighborhood-level determinants of health that influence the burden of infectious disease in early childhood (diarrhea and ARI) from a multigenerational perspective. Our specific aims include:

1. To estimate the frequency and duration of incident diarrheal disease and ARI episodes among a cohort of children ages 3-59 months, and estimate whether frequency and duration vary by exposure to intergenerational disadvantage;
2. To analyze whether healthcare utilization and treatment indicators for acute illness episodes among children ages 3-59 months vary by exposure to intergenerational disadvantage;
3. To estimate whether spatial remoteness (distance to health facilities) and/or other contextual factors moderate these relationships.

Through rigorous analysis of existing data and new measures collected through baseline interviews, daily symptom diaries, and weekly phone interviews, we seek to inform targeted public health interventions that mitigate health disparities and promote access to healthcare for young children experiencing diarrhea and ARI, two of the most common illnesses in early childhood.

## METHODS AND ANALYSIS

### Study setting

This study draws on the Chitwan Valley Family Study (CVFS). The CVFS is a general population household and community panel study, established in 1995 in western Chitwan, Nepal [71–73]. At baseline, the CVFS collected data from 151 neighborhood clusters, each including five to 15 households. Over time, the CVFS has refreshed its sample by adding additional households as they formed within or moved into sample neighborhoods. The CVFS follows all household members over time, whether or not they remain in the sample neighborhoods. This detailed prospective data collection includes measures at the individual, household, and community (neighborhood) levels, with comprehensive information describing social, demographic, and contextual data in Chitwan. These include household economic status and members’ socio-demographic characteristics, neighborhood infrastructure, and a monthly registry of household members’ location and timing of key demographic events, among many other measures. In addition, the CVFS has captured detailed measures of the location and characteristics of all health services in the western Chitwan Valley beginning in 1996, which can be linked to household and study neighborhood locations [74].

### Study design

This study includes two waves of data collection, approximately one year apart. Each wave of data collection includes a baseline interview with 52-week follow-up (Figure 1). Prior to the first wave, the sample will be randomly divided into two cohorts, or replicates, with the second replicate starting baseline interviews approximately six months after the first replicate. This ensures that respondents complete follow-up data collection throughout the calendar year even if some loss to follow up occurs. Children will be randomized to replicate 1 or 2 at the neighborhood level in order to maximize fieldwork efficiency and ensure all children in a single household are included in the same replicate.

**Figure 1.**
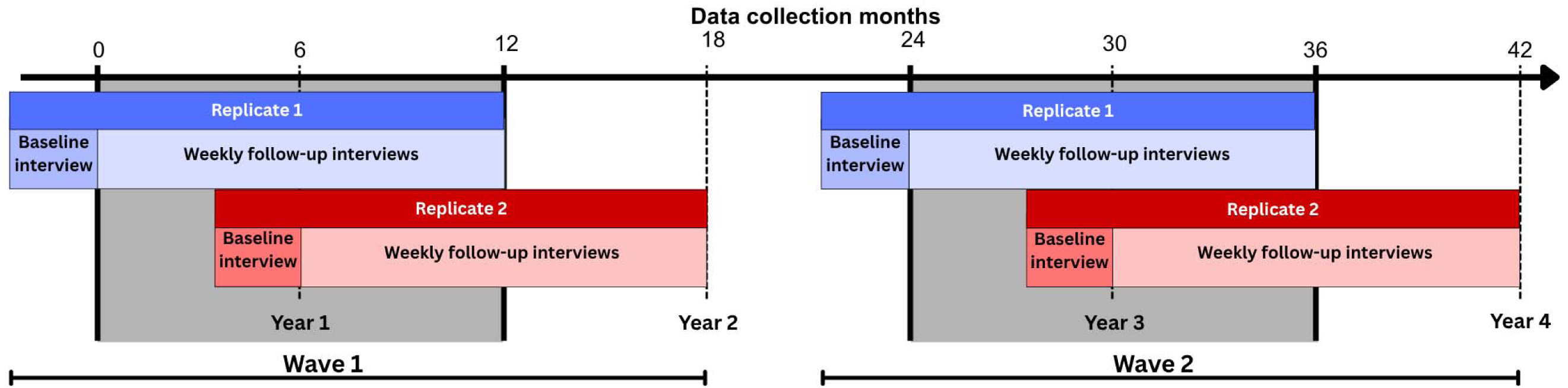
Planned data collection timeline.

The random assignment will be carried out by a study team member in the United States who is blinded to the exact neighborhood locations. Each CVFS neighborhood will be randomly assigned to replicate 1 or 2 using a randomization procedure in Stata. For children whose household is outside a CVFS neighborhood but within Chitwan, they will be assigned to the replicate corresponding to the 3-digit neighborhood code in their respondent ID, which represents the study neighborhood where they or their family member (e.g., parent, grandparent) was first enrolled in the CVFS.

### Sample and recruitment

#### Study population

The study population includes children ages 3 to 59 months at study enrollment—the baseline interview of either data collection wave—who are currently living anywhere in the western Chitwan district and co-reside with their mother. Respondents are the mothers of eligible children. Children are eligible to participate until their fifth birthday. Newly eligible children may enroll at the second wave of data collection, but no newly eligible children will enroll in either wave after baseline interviews are complete.

We will identify eligible children based on the most recent round of the CVFS household registry, the monthly demographic registry of all CVFS respondents that captures births and respondent locations. The sample will be refreshed prior to the second data collection period to include households with newly eligible children (children born since the first sample; eligible children who moved into the study area). Children who move out of the study area or reach their fifth birthday prior to the baseline interview of the second 12-month data collection period will not be included in the second wave of data collection. During both 12-month follow-up data collections, we will stop follow-up data collection for an individual child when that child ages out of eligibility or permanently moves out of the study area.

#### Inclusion/exclusion criteria

To participate in either wave, children must be (1) at least 3 months of age and less than five years of age (60 months) on the day of their baseline interview; (2) reside in the Chitwan district; (3) be currently enrolled in the CVFS; and (4) must be co-resident with their mother. Mothers must be at least 18 years of age and speak Nepali. All children who meet the inclusion criteria are eligible to participate.

Children not meeting the previously described enrollment criteria will be excluded from the study. Children will also be excluded from the study if they are five years of age or older or move out of Chitwan. Children will be excluded from the study if they have severe chronic illness or developmental delays. Only the child’s biological mother will be eligible to participate in this study. Mothers will be excluded from the interview if they have severe chronic illness or cognitive issues. Participants who move permanently outside of the Chitwan district will be excluded.

#### Sample size

Because this study relies on the existing CVFS cohort, we will include all eligible children in the sample to maximize the sample size. Across both waves, we anticipate we will enroll 979 children and their mothers (approximately 873 unique mothers). Each child will have up to 104 weeks of observations (split into two 52-week periods over the four years). Assuming a 90% response rate, which is consistent with other CVFS data collections and a 12-week pilot test [75], and participation of 715 children in each wave, we anticipate collecting up to 66,924 weekly symptom diaries (up to 15,402 child-months of observation). Given a 9.8% prevalence of diarrhea and a 6.9% prevalence of ARI in the two weeks preceding interview from a 2023 cross-sectional survey of child health indicators among children under 10 years in the CVFS, we anticipate capturing over 6,500 unique diarrhea episodes and over 4,600 ARI episodes. This diarrheal disease prevalence is similar to a nationally-representative survey in Nepal (10.4%) [76].

Table 1 shows the minimum detectable incidence rate ratio at this sample size with 80% power, alpha of 0.05, intraclass correlation of 0.10 [77], and varying prevalence of exposure to intergenerational disadvantage [Pr(X=1)] and varying coefficient of variation (CV). The power calculations are based on a two-sided ratio test of the appropriate coefficient from a generalized estimating equations (GEE) Poisson regression model that accounts for clustering of observations [78, 79].

**Table 1.**
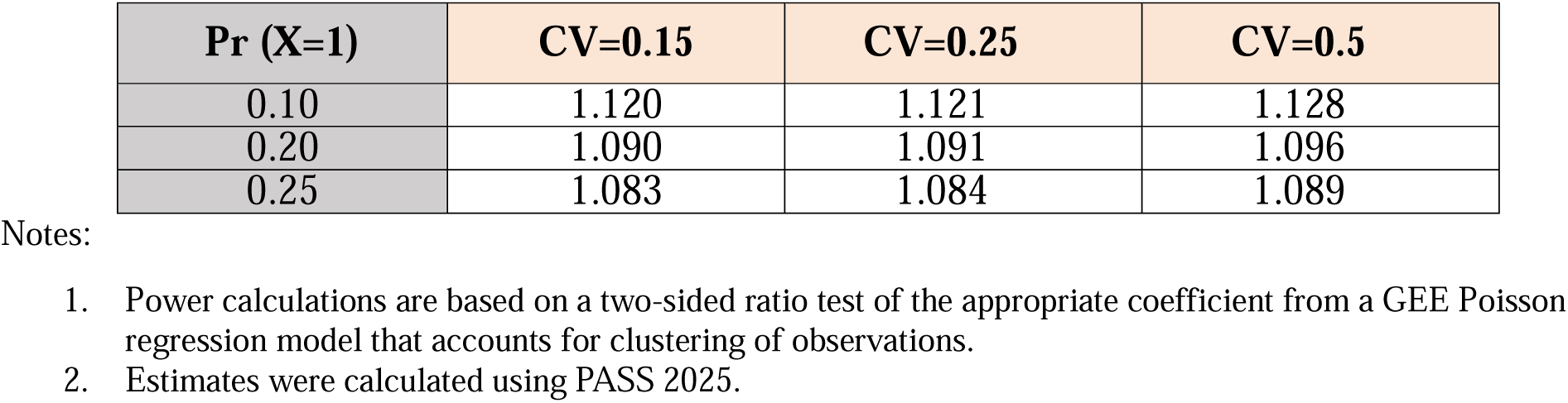
Minimum detectable incidence rate ratio (N=15,402 person-months).

| **Pr (X=1)** | **CV=0.15** | **CV=0.25** | **CV=0.5** |
| --- | --- | --- | --- |
| 0.10 | 1.120 | 1.121 | 1.128 |
| 0.20 | 1.090 | 1.091 | 1.096 |
| 0.25 | 1.083 | 1.084 | 1.089 |

Notes:
1. Power calculations are based on a two-sided ratio test of the appropriate coefficient from a GEE Poisson regression model that accounts for clustering of observations. 2. Estimates were calculated using PASS 2025.

### Data sources and collection

#### Interviewer Training

Interviewers participated in a two-week training that included general interviewing techniques and best practices in survey research, as well as study-specific content, practices, and procedures. Study-specific training prepared interviewers to complete all study procedures, including the baseline interview, the weekly follow-up phone interviews, and collection of anthropometric data from respondents and their children. Following training, interviewers trialed the baseline interview in the field for five days in neighborhoods located outside of the planned study sample. Interviewers visited households with respondents who fit study criteria (i.e., mothers with children under five years of age) and asked them if they would be willing to take part in the baseline interview process. Interviewers practiced all aspects of the baseline interview, including obtaining informed consent, completing the baseline interview, conducting household observations, and collecting anthropometric measures for mothers and their eligible child(ren).

All interviewers are native Nepali speakers. All data collection activities described in this protocol will be conducted in Nepali.

#### Baseline data collection

Prior to each baseline interview, interviewers will obtain written informed consent from the respondent. They then administer the baseline questionnaire using computer assisted personal interviewing (CAPI). The questionnaire includes modules on parent, household, and child characteristics; family dynamics and decision-making; and detailed measures of income, wealth, food security, housing characteristics, water and sanitation, indoor ventilation, and other indicators of socio-economic status. Questions about the eligible child(ren) related to their health history and daily life, including birth characteristics and location, immunization history, perceived health status, dietary diversity, stooling patterns, healthcare utilization, and childcare and school/daycare attendance. All questions in the baseline and follow-up survey instruments were translated to Nepali from English, then back-translated to ensure accuracy.

Immediately following the interviewer-administered survey, interviewers will collect anthropometric information from the respondent mother and participating child(ren) at each household. This will include height and weight data from mothers and height/length, weight, and mid-upper arm circumference from children. Interviewers use standardized equipment (height board, Seca 874U electronic scale) and adhere to international procedures to collect each measure (e.g., measuring children <24 months or <87 cm supine) [80]. The team regularly assesses instrument calibration, replaces instruments as needed, and monitors data for validity. Interviewers will record all anthropometric and immunization data on paper, which are later double-entered to ensure accuracy, and linked to study interview data.

At the conclusion of the baseline interview, interviewers will explain all procedures related to the follow-up data collection and answer any questions from respondents.

#### Follow-up data collection: Symptom diaries

Mothers will record whether each eligible child has specific symptoms each day on a printed form (diarrhea, fever, cough, runny nose, difficulty breathing or wheezing, fatigue, loss of appetite). The form is designed to be completed as a simple checklist that records whether or not a child had a specific symptom on a specific day (yes/no). The symptom diary is shown in English in Figure 2. Daily diaries are a demonstrated method for parents to record symptoms over time [81–85], even for periods as long as one year [86]. One page includes seven checklists (one week). Respondents will be given 16 weeks’ worth of printed checklists at a time, along with a pen and a waterproof storage folder. Every 4 weeks, an interviewer will collect the completed diaries, and every 16 weeks an interviewer will give the respondent a new set of printed checklists.

**Figure 2.**
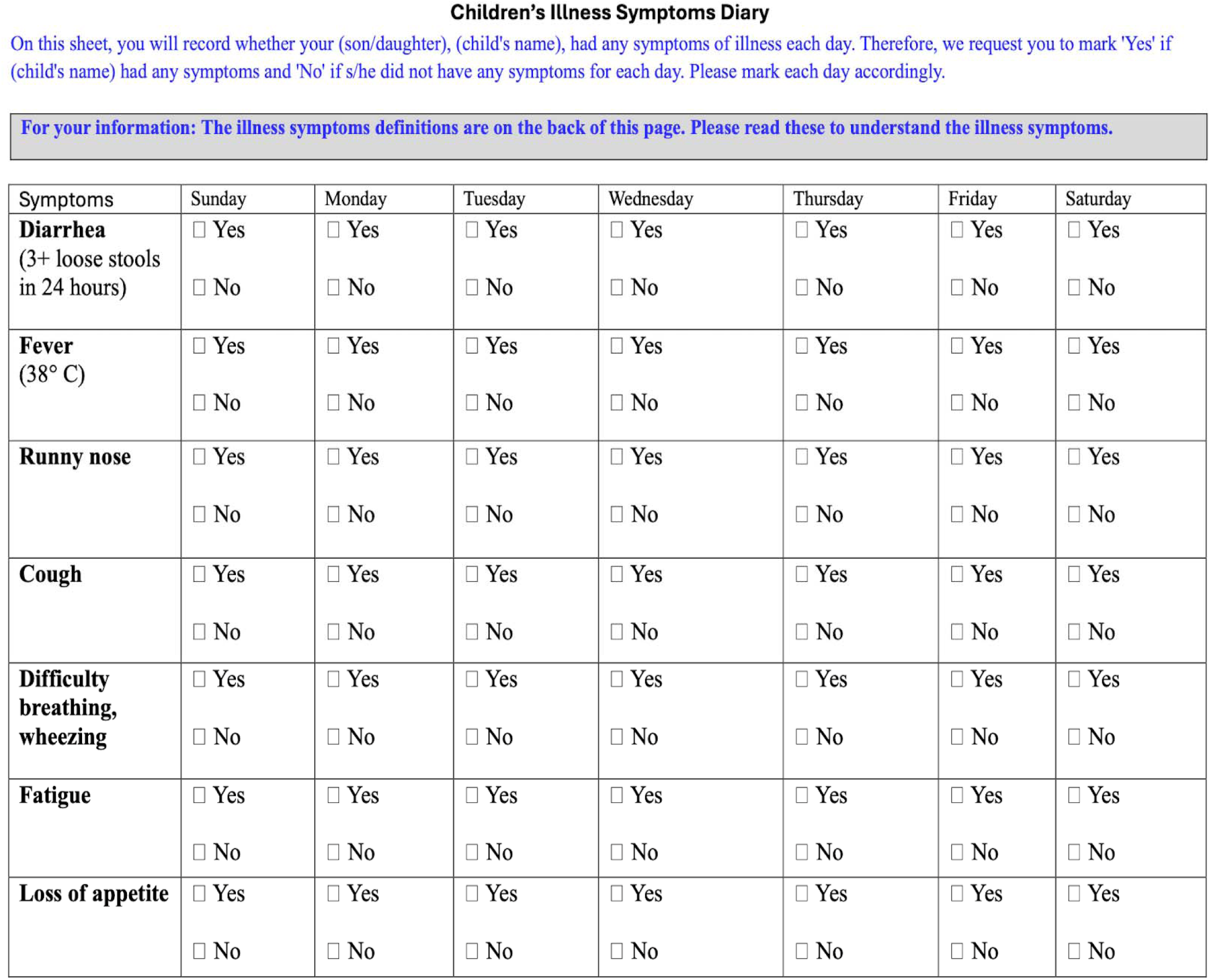
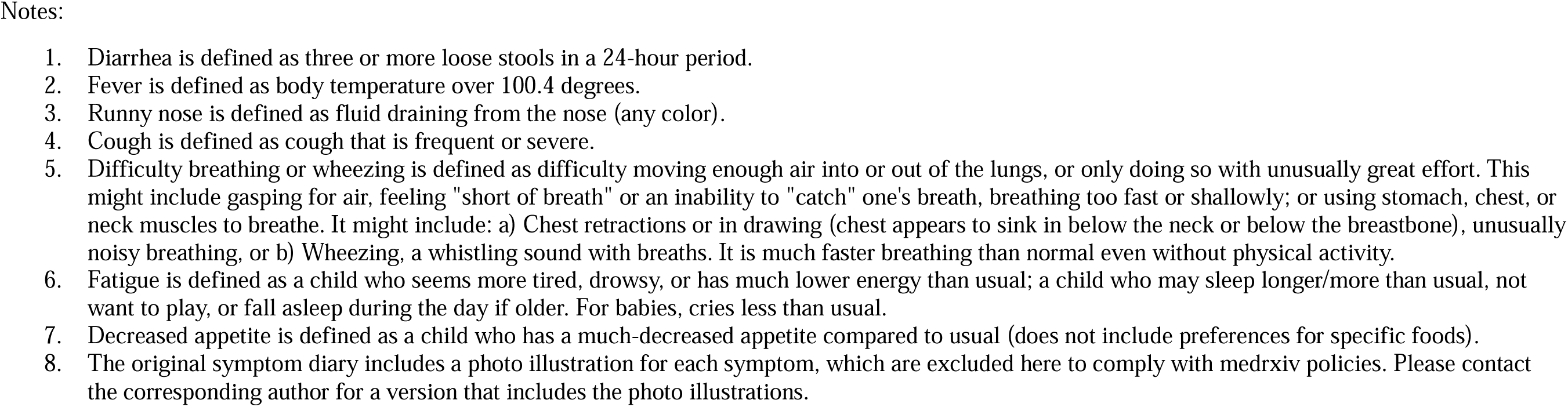
Weekly symptom diary in English.

#### Follow-up data collection: Weekly interviews

To collect the information recorded in each daily checklist, an interviewer will call each respondent at the end of each study week. During each weekly telephone interview, the interviewer will ask which symptoms each child experienced, and record which days they experienced the symptom(s). If a mother did not record daily symptoms, the interviewer will ask about the occurrence of each symptom type on each day for each child based on recall, and record that the information was provided via recall rather than daily symptom diary.

If a respondent reports a child had any symptom(s) on any day(s) in the preceding week, the interviewer will ask additional questions about whether, when, and where the child received treatment; if the child did not receive any treatment, we will ask about reasons for lack of treatment. We will ask about which family member(s) were involved in treatment decisions, expenditures related to healthcare and treatment, if the respondent or child missed work or school/daycare due to illness, and obtain names of any medication(s) administered to the child.

If a respondent cannot be reached by phone on the scheduled day, an interviewer will attempt to reach them again twice on the following day. If they cannot be reached the following day, an interviewer will attempt to interview them in person at their home two days after the scheduled day. In person interviews will use the same CAPI survey instrument as phone interviews, and will be recorded as taking place in person to allow for a control for survey mode in analyses. If a respondent is regularly unable to complete phone interviews (e.g., due to lack of phone or lack of phone in good working condition), the study team will arrange to conduct regular in person CAPI interviews on the same data collection schedule.

If a respondent cannot be reached in a study week, but is reached again the following week, we will collect the symptom checklist data for the missed week if the respondent recorded it. Missed week symptom data will be recorded by an interview using paper and pencil, then double-entered and linked to study data. We will not obtain symptom data for any missed weeks via recall nor will we collect information about healthcare utilization or treatment for symptoms reported during missed weeks due to the potential for bias.

#### Study discontinuation and loss to follow up

If a respondent cannot be reached after three phone attempts and one in-person attempt, we will not attempt to complete follow-up data collection for that week. We will resume attempting to reach the respondent again on the regularly scheduled day the following week. We will attempt to reach a respondent for four weeks, at which point we will consider them lost to follow up.

Children are no longer eligible to participate in the study once they turn five years old. Their mother will stop participating in the follow-up data collection (daily symptom checklists and weekly phone interviews) once they complete their fifth birthday. If a respondent has another eligible child, she may continue participating in the study for that child.

If a respondent and/or her children permanently move outside the study area (Chitwan district) or outside Nepal, they will no longer be eligible to participate in the follow-up data collection. In the event that a respondent or participating child dies, the family will no longer participate in the follow-up data collection.

Respondents can voluntarily withdraw from the study at any time. If a respondent misses several follow-up interviews, the interviewer will confirm they wish to continue participating at the next contact.

#### Incentives

Respondents will receive three incentives for their participation in this study: a digital thermometer, mobile phone credit, and a children’s book. At the conclusion of the baseline interview, each household will receive a digital thermometer to use throughout the study, and may keep after it ends. Mothers will receive instructions and a demonstration for its use during the baseline interview and, if the thermometer breaks during the study, the study team will replace it to ensure all participating households have a digital thermometer in good working condition to accurately assess fever.

Beginning at the baseline interview and continuing on a bi-monthly basis, mothers will receive mobile phone credit as compensation for their time completing the follow-up data collection. Finally, when a household’s participation ends due to withdrawal, after a year of data collection, when a family moves out of the study area, or on the child’s fifth birthday, the child is gifted an age-appropriate children’s book.

#### Outcomes and measures

##### Illness burden: Frequency and duration of diarrheal diseases and ARIs

We will collect information on the frequency and duration of symptoms related to diarrheal disease and ARI through daily symptom diaries and weekly phone interviews. We will leverage daily measures of symptoms to create measures of individual illness episodes. Diarrheal illness episodes will be defined as any incidence of one or more consecutive days where a child has diarrhea (3 or more loose stools in a 24-hour period). ARI episodes will be defined as the acute onset of one or more of the following respiratory symptoms: cough, runny nose, sore throat, and/or fast/labored breathing [87]. In secondary analyses, we will also create and test categorical measures of illness that capture specific constellations of symptoms, for example, diarrhea; diarrhea with fever; cough with fever; and/or others [44].

The duration of illness will be defined as the number of consecutive days a child experienced diarrhea- or ARI-related symptoms. The frequency of diarrheal disease and ARI will be defined as the number of unique illness episodes a specific child has in a defined time period (e.g., month, year).

##### Healthcare utilization outcomes

We will create a dichotomous indicator for each illness episode for whether the child utilized any healthcare services. A child will be considered to have utilized any services if he or she was evaluated by any provider, trained or untrained, at any facility outside the home or by a provider visiting the home, or the mother reported obtaining treatment from any pharmacy or drug seller, versus utilizing no healthcare services and receiving no treatment at home.

Healthcare providers with formal biomedical training (e.g., doctors, nurses, midwives, pharmacists) are more likely to correctly diagnose and treat childhood illnesses than untrained providers (e.g., drug sellers, healers, shamans) [88, 89]. A child will be considered to have utilized care with a trained provider if he or she utilized care at a primary, secondary, or tertiary health facility, a pharmacy, or was evaluated by a doctor, nurse, other healthcare worker, or pharmacist; versus care with or evaluation by an untrained provider (drug shop, drug seller, other provider) or no care.

We will create continuous measures of the timing of the first healthcare utilized and the first trained provider utilized relative to symptom onset (days), and a dichotomous measure of whether the child utilized a health service within 24 hours of symptom onset.

Leveraging the detailed data we will collect on healthcare utilization for each illness episode, we will create utilization indicators related to other aspects of health services (e.g., facility characteristics, such as whether a facility has x-ray; open hours, such as whether a facility is open 24 hours and/or seven days per week; level of care, such as whether the child attended a secondary facility; sector of care, such as whether the child attended a private sector facility) and the number of providers/facilities visited. We will also create measures related to whether the child experienced barriers to care and whether the family incurred debt for care-related expenditures.

Finally, in the weekly interview, we will collect data on the type of medication(s) administered to a child in each illness episode; if we obtain sufficient sample sizes and details, we will assess whether a child was administered an appropriate medication for the specific symptoms reported.

##### Intergenerational disadvantage

Our primary exposure of interest is intergenerational disadvantage, that is, experiences of low SES among two or more generations in the household. To operationalize this exposure, we will create a composite measure of intergenerational disadvantage based on the following indicators: (1) household wealth and income; (2) employment; and (3) social group (ethno-caste group). This measure will draw on data on children’s current household circumstances, as well as measures of SES and household characteristics prospectively measured during parent’s childhoods in prior CVFS data collections. For parents whose household was not enrolled in the CVFS during their childhood, we will draw on retrospective measures of childhood experiences and household circumstances collected during the current data collection. We will collect measures of income, employment, as well as physical housing characteristics and durable goods ownership to estimate wealth quintiles [90]. Within-study wealth quintiles can be compared to nationally-representative samples to describe relative wealth status [91].

Disadvantage is a multidimensional construct. As such, as we will also operationalize and test indicators of economic and social disadvantage separately. Households will be considered economically disadvantaged if they are in the poorest wealth quintile when standardized to the nationally representative DHS sample; if they report food insecurity; if their household is below the national poverty line; or if a household member currently engages in wage labor. If there is a lack of overlap in households identified using these measures, we will develop a contextually-appropriate composite measure based on the Nepal Multidimensional Poverty Index [92]. We will assess an indicator of household members’ social group, which is a setting-specific indicator of SES.

Children will be considered exposed to intergenerational disadvantage if their household is currently disadvantaged, and one or both of their parents experienced disadvantage in their childhood, as defined by meeting one or more of the above criteria. We will also test models with indicators of current SES, comparing children in currently disadvantaged households to those who are not currently disadvantaged, as well as models with indicators comparing children whose parent(s) experienced disadvantage to those who did not, stratified by current SES. The latter will allow us to isolate the effect of disadvantage in the prior generation on children’s outcomes among those who are not currently disadvantaged.

##### Distance to health facilities and neighborhood contextual measures

The CVFS includes precise GIS-based location information for each of the 151 neighborhoods included in the study (neighborhood centroid), household location for any CVFS households in western Chitwan that are outside the boundaries of study neighborhoods, as well as GIS locations of all health facilities included in prior health service history calendars (1996, 2008, 2016; HHC). We will obtain GIS data for any new health services in updated HHCs during this data collection.

To estimate distances to health facilities, we will generate a dataset that includes the GIS location of each neighborhood and each health service in the Chitwan Valley. We will estimate distance from each neighborhood to (1) the nearest health service (any); (2) the nearest health facility staffed by a trained provider (e.g., nearest public- or private-sector primary clinic, hospital; trained providers include doctors, nurses, and midwives); and (3) the nearest secondary or tertiary facility (e.g., hospital). These include great-circle distance (orthodromic distance), which is the shortest distance between two points on a sphere; road distance, leveraging existing spatial data from the Global Roads Database, which can be conceptualized as travel distance or travel time [93]; and cost-based distance, which accounts for physical hazards and slope in the distance between two locations. We will also generate a matrix of the great-circle and cost-based distances from each household/neighborhood centroid to each health facility in the HHC.

Because most CVFS neighborhoods are relatively small in geographic scale, a neighborhood centroid is a reasonable proxy. However, if household-level GIS measures for household within neighborhoods become available over the course of this study, we will use the more precise household-level GIS measure for all households.

To determine concentration of disadvantage in a neighborhood, we will assess variation in the counts and proportions of all households in the neighborhood that meet the definition of intergenerational disadvantage. We will draw on existing approaches to assess neighborhood disadvantage using composite approaches [94, 95]. In selecting measures, we will draw on the large literature examining variation in characteristics across neighborhoods and/or census tracts in the US, testing models that use crude means, quantiles, and spatial empirical Bayes approaches (e.g., [96–99]).

#### Statistical analysis

We will estimate the incidence of diarrhea and ARI based on the number of unique illness episodes per 1,000 children at risk (i.e., children under five years completing symptom diaries in the same period). We will also estimate the incidence of the more specific symptom combinations using the categorical variable described above, following the same procedure. We will estimate the frequency of diarrhea and ARI by estimating the incidence rates of diarrhea and ARI using a Poisson distribution [100]. We will estimate mean duration of illness as the mean number of days of symptoms for diarrhea and ARI illness episodes for each child. We will descriptively assess whether the incidence and duration of diarrhea and ARI illness episodes varies among children exposed versus unexposed to intergenerational disadvantage, as well as among those currently disadvantaged versus not currently disadvantaged. We will use other descriptive approaches such as sequence plots to characterize illness and healthcare utilization trajectories over time.

We will then estimate the association between intergenerational disadvantage and illness frequency and duration using mixed-effects regression models that account for clustering in the data via child-level random effects, and control for child and household characteristics. We will also test models that include household and/or neighborhood level random effects. In addition to the mixed-effects regression models, we will estimate more flexible models that include robust standard errors clustered at the neighborhood level, and generalized linear regression models with GEE to obtain population-averaged estimates. For all analyses, we will test unadjusted models and models that adjust for the child’s age and sex. We will also test models that adjust for birthweight as a potential indicator of underlying health conditions. For each analysis testing for differences in outcome by exposure to intergenerational disadvantage, we will test a separate model for the composite measure of intergenerational disadvantage, as well as each of the three individual dimensions of intergenerational disadvantage (wealth and income, employment, social group).

Next, we will estimate relationships between illness burden, healthcare utilization, and intergenerational disadvantage. We will use a similar modeling strategy as described above--mixed-effects regression models with the appropriate link function, followed by models with clustered standard errors and GEE models—to estimate the associations between ARI and diarrhea frequency and duration and healthcare utilization outcomes, followed by estimation of the association between intergenerational disadvantage and healthcare utilization. To test for effect modification of illness frequency and duration by exposure intergenerational disadvantage, we will add an interaction term to the mixed-effects regression models.

Drawing on our GIS data, we will conduct descriptive analyses of the spatial distribution and clustering of key exposure (intergenerational, current disadvantage) and outcome measures (illness frequency, duration, healthcare utilization). To examine clustering, we will generate choropleth maps that describe the degree of clustering per neighborhood. We will also estimate spatial autocorrelation through the use of statistical tests such as Moran’s *i.* To estimate the associations between health facility distance and outcomes, we will include continuous measures of facility distance in the mixed-effect regression models. We will estimate associations between an indicator of neighborhood concentration of disadvantage and the outcomes of interest, then test for modifying effects of neighborhood disadvantage by adding an interaction term for distance or concentration of disadvantage to the mixed-effects regression models specified above. Finally, we will also test for modifying effects of facility distance by adding an interaction term for facility distance to the mixed-effects regression models specified above.

Given the number of primary exposure-outcome models being tested, we will apply the Benjamini-Hochberg procedure [101] to control the false discovery rate at 0.05, rather than a Bonferroni-type correction, given the correlated nature of our outcomes and exposure measures and to avoid an overly conservative reduction in power to detect true associations.

We will assess to what extent data are missing due to loss to follow up, and will assess missing data for non-attrited observations. We will assess whether and how each type of missingness is associated with the exposure(s) of interest. In sensitivity analyses, we will use complete case analysis, testing models where (1) any children who are lost to follow up are excluded; and (2) only observations without missing covariate data are included.

### Patient and Public Involvement

While patients and the broader public were not directly involved in the development of this study protocol, CVFS respondents helped shaped the research questions based on their stated priorities related to children’s health research in prior data collections. Once the study is completed, results will be disseminated to participants via a research brief. We will also hold dissemination workshops for local stakeholders in Chitwan (e.g., healthcare workers, health facility managers, local and district government) and at the national level.

## ETHICS AND DISSEMINATION

### Ethical and safety considerations

We received IRB approval from the Nepal Health Research Council (Registration number 124_2025) and the University of Michigan (HUM00260534).

Surveyors will obtain informed consent from all respondents prior to enrollment in the study, including for the baseline interview, follow-up data collection, and for their children’s participation in anthropometric data collection. Identifying information (i.e., respondent name, phone number) will be stored separately from the survey data, linked by unique study ID. Household and neighborhood GIS data is also stored separately from identifying information Access to identifying information will be restricted to the data collection team in Nepal. All other study team members will access only de-identified data. These practices build on the CVFS’ long history of successfully protecting respondent data and privacy.

In the case that a respondent reports that her child experienced a danger sign as classified by the WHO/UNICEF Integrated Management of Childhood Illness clinical guidelines used in Nepal [102], such as bloody stools or difficulty breathing, the interviewers will assess whether the child has sought appropriate treatment. For this study, appropriate healthcare options for danger signs include hospitals, health posts, health centers, private clinics, or non-governmental organization sector facilities, which are all staffed by trained nurses and/or doctors. Female community health volunteers, pharmacies, drug shops, traditional or religious practitioners, or home treatment are not considered adequate for danger signs. If the child has not received sufficient treatment for the symptoms reported (e.g., reported a danger sign but no healthcare was sought outside the home), interviewers are trained to report this to their supervisor immediately following the conclusion of the interview. Supervisors will alert a study nurse, who will reach out to each family and provide advice about symptom management and where to seek medical care.

In the case that a study respondent has a question about medical treatment or any other medical topic, interviewers will never answer these questions themselves, but rather refer them to the study nurse. The interviewer will record the question and let the respondent know that a study nurse will contact them to respond to their question.

### Dissemination

Study results and findings will be shared through conferences and published in peer-reviewed journals following the International Committee of Medical Journal Editors guidelines. Findings will also be disseminated via workshops with loca1, national, and international stakeholders in pediatric healthcare delivery including healthcare workers, local and national policymakers, public health practitioners, and other relevant stakeholders.

We will disseminate comprehensive, well-documented public and restricted-use data through the Data Sharing for Demographic Research (DSDR) program at the Inter-university Consortium for Political and Social Research (ICPSR) at the University of Michigan. Beyond making the data and comprehensive supporting documentation freely available to users globally, disseminating new data products is core to the success of enabling secondary analyses by the scientific and public health communities. To increase the impact of this unique data resource, we will promote the data release via the CVFS website, publications, listservs, social media, and dissemination workshops. The CVFS website has detailed information to support data use, including the sample and methods for each unique data collection, interviewer procedures, response rates, content, directions for merging with other datasets, publications using the data, links to the full codebooks, and data access instructions including links to the ICPSR/DSDR webpage. All survey instruments will be available in English and Nepali.

## Discussion

This protocol describes a prospective, multigenerational cohort study that will leverage the unique infrastructure of the CVFS to examine whether and how intergenerational disadvantage shapes the burden of diarrheal disease and ARI in early childhood, and the ways in which families respond to these illnesses over time. By combining high-frequency symptom surveillance—through daily diaries and weekly interviews over two 52-week periods—with over three decades of existing individual, household, and neighborhood data, this study is uniquely positioned to address critical gaps in understanding the social patterning of childhood infectious disease morbidity and healthcare utilization.

Expected contributions include rigorous estimates of diarrhea and ARI incidence in this population; novel evidence on whether persistent, multigenerational socioeconomic disadvantage confers risks beyond those captured by contemporaneous socioeconomic measures; detailed characterization of treatment-seeking trajectories during specific illness episodes and across early childhood; and assessment of how spatial and contextual factors, including distance to health facilities and neighborhood concentration of disadvantage, moderate these relationships. Findings from this study are expected to inform the design of targeted public health interventions and health system strengthening efforts that address the compounding effects of intergenerational disadvantage on child health, with implications for similar settings in Nepal and other low- and middle-income countries where diarrheal disease and ARI remain leading causes of under-five morbidity and mortality.

## Data Availability

Data have not yet been generated for this study. We will disseminate public and restricted-use data through the Data Sharing for Demographic Research (DSDR) program at the Inter-university Consortium for Political and Social Research (ICPSR) at the University of Michigan.

## List of abbreviations

ARI: Acute respiratory infection
CV: Coefficient of Variation
CVFS: Chitwan Valley Family Study
DSDR: Data Sharing for Demographic Research
GEE: Generalized estimating equation
HHC: Health history calendar
ICPSR: Inter-university Consortium for Political and Social Research
SES: Socio-economic status

## Declarations

## Author Contributions

The study was conceptualized by ET, GN, and JK, with input from DG, IC, RG, and WGA. ET, IC, and MD drafted the original study protocol with input from RG, DG, AS, AH, and YS. ET and JK conducted the power calculations. ET and MD prepared the protocol manuscript. All authors reviewed and provided feedback on the final version of the manuscript.

## Acknowledgements

We are grateful to staff at the Institute for Social Research—Nepal for support in the data collection and related tasks. Krishna Shrestha supported programming of data collection instruments for the pilot and current data collections. Gurung and Deepika Bagale provided significant assistance with data management. Bluvision Subedi and Dil Bahadur C.K. supported data collection, and we are grateful to the study interviewers. Heather Gatny and Jennifer Mamer provided additional research support.

## Data Statement

We will disseminate public and restricted-use data through the Data Sharing for Demographic Research (DSDR) program at the Inter-university Consortium for Political and Social Research (ICPSR) at the University of Michigan.

